# No Heavy Drinking Days and Percentage of Days Abstinent in a Randomized Trial of Adjunctive Ketamine for Alcohol Use Disorder: Secondary Analysis of the KARE Trial

**DOI:** 10.64898/2026.09.26.26363761

**Authors:** Celia JA Morgan, David J. Nutt

## Abstract

In the KARE trial, 96 adults with severe alcohol use disorder, abstinent following detoxification, were randomised in a 2×2 factorial design to 3 weekly infusions of ketamine (0.8 mg/kg) or saline, each combined with mindfulness-based psychological support (PS) or alcohol psychoeducation (PE). We re-expressed the trial data on endpoints acceptable to regulators and compared the most and least intensive arms. Across pooled weeks 17–24, ketamine plus PS gave 78.5% versus 57.8% days abstinent (difference, 20.7 percentage points; P = .032); adjusted for baseline, the difference at weeks 21–24 was 26.9 percentage points (P = .011). At weeks 21–24, 50.0% versus 26.1% had no heavy drinking days (P = .127). Relative reductions in heavy drinking days were 55% in KARE, against 25%, −17%, 46% and 35% reported for naltrexone, exenatide, semaglutide and pemvidutide. These post hoc differences, measured 4 to 6 months after a 4 week course ended, are of magnitude similar to agents requiring continuous administration.

## Introduction

Glucagon-like peptide-1 (GLP-1) receptor agonists have attracted intense interest as treatments for alcohol use disorder (AUD), on a randomized evidence base that is developing and not uniform: exenatide did not separate from placebo,^1^ whereas semaglutide has done so in two trials, the larger restricted to patients with comorbid obesity.^2,3^

How effect sizes from this class compare with another novel approach, ketamine therapy, is unexamined. We re-expressed KARE trial data^4^ on endpoints acceptable to regulators^5-7^ and set both alongside an approved treatment, depot naltrexone,^8^ and the GLP-1/glucagon co-agonist pemvidutide, reported in a press release (RECLAIM).^9^

## Methods

KARE randomised 96 adults with severe AUD, abstinent after detoxification, to 3 weekly infusions of ketamine (0.8 mg/kg) or saline, each with 7 sessions of mindfulness-based relapse prevention psychological support (PS) or alcohol psychoeducation (PE). Participants kept daily drinking diaries for 6 months. The trial was approved by the South West–Central Bristol Research Ethics Committee (15/SW/0312); participants gave written informed consent.

An HDD was defined as ≥8.75 UK units (70 g alcohol) for men and ≥7.0 units (56 g alcohol) for women. Outcomes were assessed over 28-day windows at weeks 17–20 and 21–24 and pooled weeks 17–24: the proportion with no HDD, the percentage of days abstinent, and abstinence throughout days 1–154. Baseline abstinence came from a 2-week pre-treatment record. Analyses were observed-case. This analysis addresses the active combination against the least intensive comparator, ketamine plus PS versus placebo plus PE. Tests were Welch t tests, Fisher exact tests, and analysis of covariance adjusted for baseline. All analyses are post hoc, with nominal 2-sided p values unadjusted for multiplicity.

Comparator data came from published reports of naltrexone,^8^ exenatide^1^ and semaglutide^2,3^ and from the topline report of pemvidutide.^9^ Where absolute drinking was not stated, rates were derived from reported baselines and change scores, assuming for pemvidutide a baseline of 7 heavy drinking days per week.

## Results

Of 96 randomized, 85 provided diary data in the follow-up windows. Drinking fell in every arm: mean days abstinent rose from 3.4% pretreatment to 70.0% at weeks 17–24, including on placebo plus PE (4.0% to 57.8%) (Table 1).

**Table 1.** Abstinence and Absence of Heavy Drinking, Ketamine Plus PS vs Placebo Plus PE.

|  | Ketamine + PS<br>(n=20) | Placebo + PE<br>(n=23) | P value<sup>a</sup> |
| --- | --- | --- | --- |
| **Days abstinent, %** |  |  |  |
| Baseline (pretreatment) | 5.2 | 4.0 | — |
| Weeks 17–20, mean (SD) | 79.1 (28.2) | 58.5 (35.0) | .039 |
| Weeks 21–24, mean (SD) | 77.7 (26.6) | 57.1 (37.2) | .041 |
| Weeks 17–24 pooled, mean | 78.5 | 57.8 | .032 |
| **No heavy drinking days, No. (%)** |  |  |  |
| Weeks 17–20 | 12 (60.0) | 7 (30.4) | .069 |
| Weeks 21–24 | 10 (50.0) | 6 (26.1) | .127 |
| Abstinent throughout days 1–154, No. (%) | 7 (35.0) | 5 (21.7) | .497 |

Abbreviations: PE, psychoeducation; PS, psychological support. Percentages are of observed diary days. The 2 intermediate arms of the factorial design (ketamine plus PE, placebo plus PS) are not shown; all 4 arms appear in the primary publication.<sup>4</sup> <sup>a</sup>P values are from Welch t tests for means and Fisher exact tests for proportions, are nominal, and are unadjusted for multiplicity.

Ketamine plus PS gave a higher percentage of days abstinent at every window: 79.1% vs 58.5% at weeks 17–20 (difference, 20.6 percentage points [95% CI, 1.1-40.0]; P□=□.039), 77.7% vs 57.1% at weeks 21–24 (20.6 [0.9-40.4]; P□=□.041), and 78.5% vs 57.8% pooled (20.7 [1.9-39.5]; P□=□.032). Adjusted for baseline, the pooled difference was 26.2 percentage points (6.6-45.7; P□=□.010).

More had no HDD, though not significantly: 60.0% vs 30.4% at weeks 17–20 (odds ratio, 3.43 [95% CI, 0.97-12.09]; P□=□.069) and 50.0% vs 26.1% at weeks 21–24 (2.83 [0.79-10.17]; P□=□.127). Abstinence throughout days 1–154 reached 35.0% vs 21.7% (1.94 [0.50-7.49]; P□=□.497).

Across pooled weeks 17–24, HDD comprised 10.1% vs 22.2% of days, a 55% relative reduction (P□=□.062); values were 7.3% vs 22.2% at weeks 17–20 (67% reduction; P□=□.018) and 13.0% vs 22.0% at weeks 21–24 (P□=□.226). Comparator reductions are shown in Figure 1.

**Figure 1.**
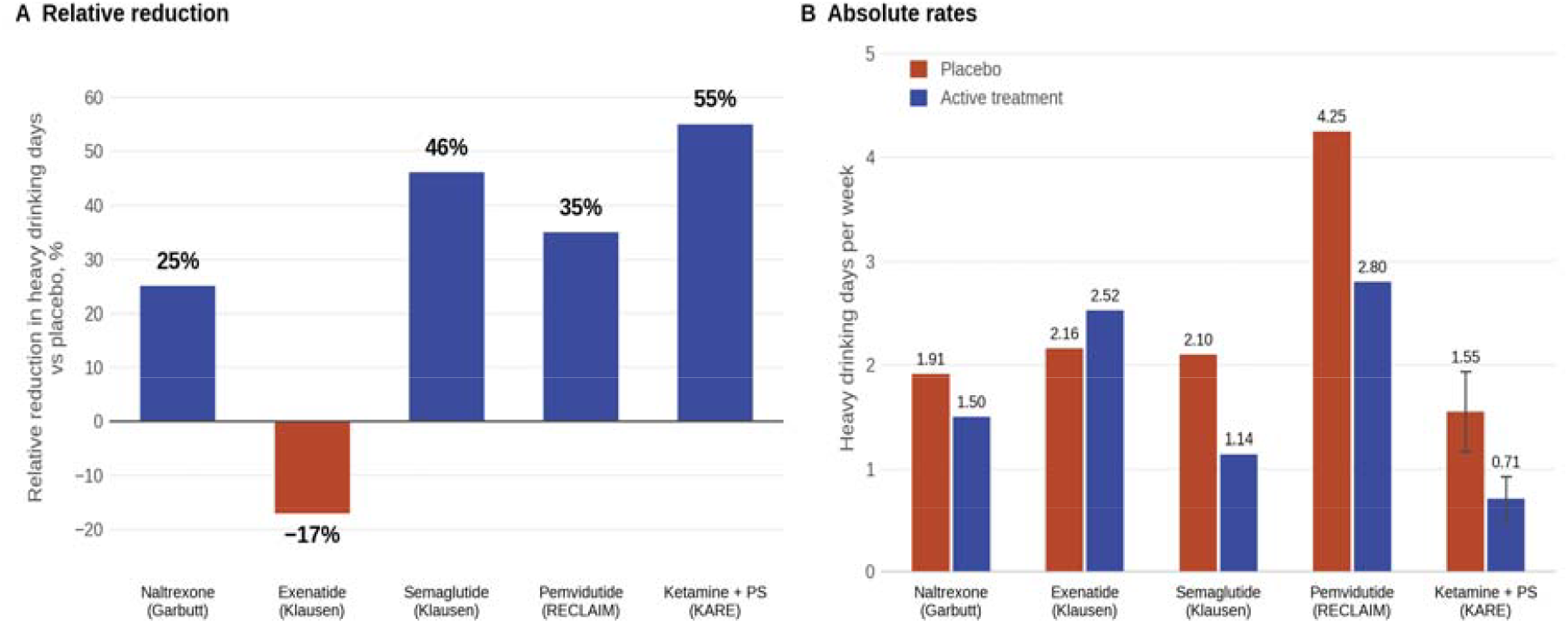
Heavy Drinking Days in KARE and in Trials of Established and Emerging Pharmacotherapies. A, Relative reduction in heavy drinking days versus placebo, calculated as 1 − (active rate ÷ placebo rate). B, Absolute heavy drinking days per week in each arm, showing the differing baseline severity that makes the relative reductions in A only indirectly comparable. KARE values compare ketamine plus PS with placebo plus PE over pooled weeks 17–24; error bars are 1 SE. Relative reductions were 25% (naltrexone), −17% (exenatide), 46% (semaglutide) and 35% (pemvidutide). Comparator values are derived from published baselines and change scores; for pemvidutide a baseline of 7 heavy drinking days per week was assumed (see Methods). Populations, follow-up and heavy-drinking-day thresholds differ between trials and these comparisons are indirect

## Discussion

Ketamine with psychological support was associated with approximately 21 more percentage points of days abstinent than the placebo plus PE arm, 4 to 6 months after the final infusion. Differences in the proportion with no HDD were in the same direction but not significant. The contrast is between complete treatment packages, and the separate contributions of ketamine and psychological support cannot be distinguished within it.

The cross-trial comparison requires caution. Participants entered KARE detoxified, so improvement occurred in every arm, and placebo-arm heavy drinking ranged from 1.6 days per week in KARE to an estimated 4.3 in RECLAIM. Absolute rates are therefore not comparable, and relative reductions are the more interpretable measure.

Limitations include small samples, post-hoc analysis and reliance on a company topline report for one comparator and differences in patient characteristics across samples. KARE used a 2×2 factorial design, and the contrast reported here does not estimate the main effects of ketamine or of psychological support, which appear in the primary publication;^4^ the contribution of each component cannot be determined from this analysis. Adequately powered evaluation is underway in the MOREKARE phase 3 trial.

The enthusiasm around incretin agonists concerns treatments given indefinitely, with effect dependent on continued exposure. The differences here were measured 4 to 6 months after a 4-week course ended. If effect sizes across mechanisms prove broadly similar, the more discriminating comparisons may be the persistence of benefit once treatment stops and the burden required to sustain it, properties on which brief, time-limited interventions warrant the attention currently directed at continuous pharmacotherapy.

## Data Availability

Deidentified participant-level data, the analysis code used for this secondary analysis, and the data dictionary will be made available to investigators whose proposed use of the data has been approved by an independent review committee, following publication and on receipt of a signed data access agreement. Requests should be directed to the corresponding author.

## Article Information

### Trial Registration

ClinicalTrials.gov NCT02649231.

### Author Contributions

Dr Morgan had full access to all of the data in the study and takes responsibility for the integrity of the data and the accuracy of the data analysis.

### Concept and design

CJAM, DJN.

### Acquisition, analysis, or interpretation of data

CJAM, DJN.

### Drafting of the manuscript

CJAM.

### Critical review of the manuscript for important intellectual content

DJN.

### Statistical analysis

CJAM.

### Obtained funding

CJAM.

